# The epidemiological characteristics of stroke phenotypes defined with ICD-10 and free-text: a cohort study linked to electronic health records

**DOI:** 10.1101/2023.04.03.23288096

**Authors:** Emma M Davidson, Arlene Casey, Claire Grover, Beatrice Alex, Honghan Wu, Archie Campbell, Fionna Chalmers, Mark Adams, Matthew Iveson, Andrew M McIntosh, Emily Ball, Kristiina Rannikmae, Heather Whalley, William N Whiteley

## Abstract

**Background:** Coded healthcare data may not capture all stroke cases and has limited accuracy for stroke subtypes. We sought to determine the incremental value of adding natural language processing (NLP) of free-text radiology reports to international classification of disease (ICD-10) codes to phenotype stroke, and stroke subtypes, in routinely collected healthcare datasets.

**Methods:** We linked participants in a community-based prospective cohort study, Generation Scotland, to clinical brain imaging reports (2008-2020) from five Scottish health boards. We used five combinations of NLP outputs and ICD-10 codes to define stroke phenotypes. With these phenotype models we measured the: stroke incidence standardised to a European Standardised Population; adjusted hazard ratio (aHR) of baseline hypertension for later stroke; and proportion of participants allocated stroke subtypes.

**Results:** Of 19,026 participants, over a mean follow-up of 10.2 years, 1938 had 3493 brain scans. Any stroke was identified in 534 participants: 319 with NLP alone, 59 with ICD-10 codes alone and 156 with both ICD-10 codes and an NLP report consistent with stroke. The stroke aHR for baseline hypertension was 1.47 (95%CI: 1.12-1.92) for NLP-defined stroke only; 1.57 (95%CI: 1.18-2.10) for ICD-10 defined stroke only; and 1.81 (95%CI: 1.20-2.72) for cases with ICD 10 stroke codes and NLP stroke phenotypes. The age-standardised incidence of stroke for these phenotype models was 1.35, 1.34, and 0.65 per 1000 person years, respectively. The proportion of strokes not subtyped was 26% (57/215) using only ICD-10, 9% (42/467) using only NLP, and 12% (65/534) using both NLP and ICD-10.

**Conclusions:** Addition of NLP derived phenotypes to ICD-10 stroke codes identified approximately 2.5 times more stroke cases and greatly increased the proportion with subtyping. The phenotype model using ICD 10 stroke codes and NLP stroke phenotypes had the strongest association with baseline hypertension. This information is relevant to large cohort studies and clinical trials that use routine electronic health records for outcome ascertainment.

## Introduction

Some health outcomes are poorly characterised in routinely collected data, and are difficult to use for research and healthcare improvement. For example, although data coded with the International Classification of Disease 10 (ICD-10) identify cases of all-stroke (hemorrhagic, ischemic and unspecified stroke combined), these data often lack stroke subtyping.^1^ Stroke phenotypes in radiologists’ reports of brain images could be extracted with natural language processing (NLP), an automated method to examine large amounts of free-text,^2-6^ and thereby provide an opportunity to improve subtyping of strokes.

Definitions of disease phenotypes (phenotype models) in electronic health records (EHR) increasingly include both simple data (e.g. code lists) and more complex data elements, such as NLP of free-text. ^7, 8^ To encourage a structured approach to documenting these phenotype models, ‘computable phenotypes’ have been developed to provide a machine-processable representation of these combined data elements ^9, 10^ Computable phenotypes require cycles of testing, refinement and validation. ^7^

For this purpose, we developed five different phenotype models for stroke: one using NLP of brain image reports only, one with hospital admission coded data (ICD-10) only, and three phenotype models combing both NLP of brain image reports and coded data in different ways. We tested these phenotype models with data from participants in a community based cohort study, Generation Scotland (GS), linked to EHR and brain imaging reports, by estimating the following: (i) all-stroke incidence; (ii) the association between baseline diagnoses of hypertension and all-stroke outcome; and (iii) the proportion of patients with unclassified stroke subtype.

## Methods

### Data sources

#### Generation Scotland

GS is a cohort of approximately 24,000 people who were recruited, across Scotland, for the GS 21st Century Genetic Health study (21CGH) and the GS Scottish Family Health Study (SFHS). 21CGH was created to look at the genetic profile of a control population living in Scotland in relation to health and disease, with all participants having at least three grandparents born in Scotland.^11^ Recruitment took place in six different locations across Scotland (Aberdeen, Banff, Dundee, Edinburgh, Glasgow and Peterhead) from 2007-2009 and there were nearly 2,000 participants.^12^ SFHS was a much larger family-based epidemiology study that recruited about 24,000 participants across Scotland from 2006-2011, and included 5573 family groups (excluding people with no relations involved). Upon recruitment, GS participants had baseline data collected including physical measures (e.g. blood pressure), health data (e.g. past medical history) and biological samples.^12, 13^ Participants also gave permission for the use of their health data for all approved health related purposes and, therefore, data of any health events following recruitment can be obtained from routinely coded healthcare datasets (e.g. Scottish Morbidity Record (SMR)). We collaborated with GS to obtain, and apply NLP to, the reports of any clinical brain imaging undergone by GS participants following their recruitment to GS (2006-2011) until the end of 2020.

#### Brain imaging reports and NLP outputs

We obtained 8511 brain imaging reports from five Scottish health boards: Tayside, Fife, Grampian, Lothian and Greater Glasgow & Clyde. Scan reports, pseudonymised with GS study identification numbers (GSIDs), were sent to GS, and stored in their secure server. Reports (2931) were excluded for the following reasons: research scans (not carried out as part of routine clinical care), not CT or MRI scans, no content, only clinical history details and scans of other body parts (Supplementary figure 1).

We used EdIE-R ^2^ to extract brain phenotypes from the free-text brain imaging reports. EdIE-R is a rule-based NLP system which identifies 24 cerebrovascular and other neurological phenotypes: ischemic stroke (deep recent, deep old, cortical recent, cortical old and underspecified); hemorrhagic stroke (deep recent, deep old, lobar recent, lobar old and underspecified); stroke underspecified, tumour (meningioma, metastasis, glioma and other); small vessel disease; atrophy; subdural haematoma; subarachnoid hemorrhage (aneurysmal and other); microbleed (deep, lobar and underspecified); hemorrhagic transformation. When tested on NHS Tayside routinely collected brain scans, EdIE-R had a positive predictive value (PPV) and sensitivity for expert report reading of: cerebral small vessel disease, 100% and 100%; any ischemic stroke, 85% and 89%; and hemorrhagic stroke, 72% and 96%. ^14^ When tested on the GS dataset, EdIE-R had an F1 score against annotated reports of 93% for ischemic stroke (PPV 88% and sensitivity 98%).^15^

#### GS baseline and outcome data

Participants’ baseline data were smoking status; self-reported history of stroke, cardiovascular disease, diabetes and depression; and measurements of blood pressure (BP) and body mass index (BMI). We excluded 2087 scans belonging to participants whose data could not be linked due to missing or incorrect ID information, whose linked health board was not one of the five we received scans from or who had any of the aforementioned baseline variables missing (Supplementary figure 1). Outcome data consisted of routinely coded hospital admission data (general admissions; SMR01), and was defined as ischemic stroke (ICD-10 I63), hemorrhagic stroke (ICD-10 I61) and unspecified stroke (ICD-10 I64). GS also provided the dates of any participants withdrawing from the study or dying during follow up so that we could right censor these data.

### Population

Our study population was GS participants normally resident in the Scottish health boards of Tayside, Fife, Grampian, Lothian and Greater Glasgow & Clyde with complete baseline information, followed up from joining GS (2006-2011) to the end of 2020.

### Definition of phenotype models

We defined five different phenotype models for stroke using the NLP output and coded data (Table 1).

**Table 1:**
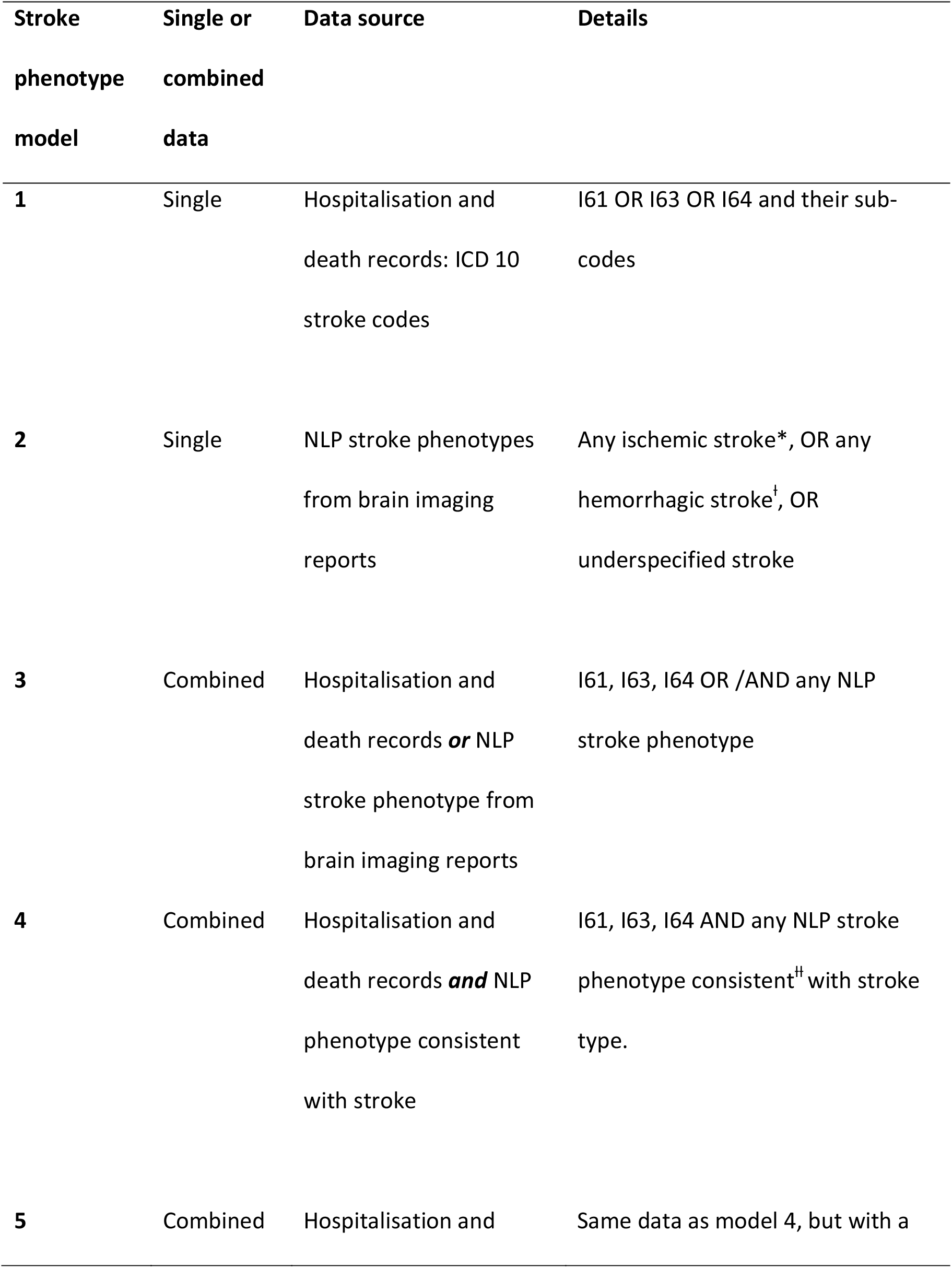
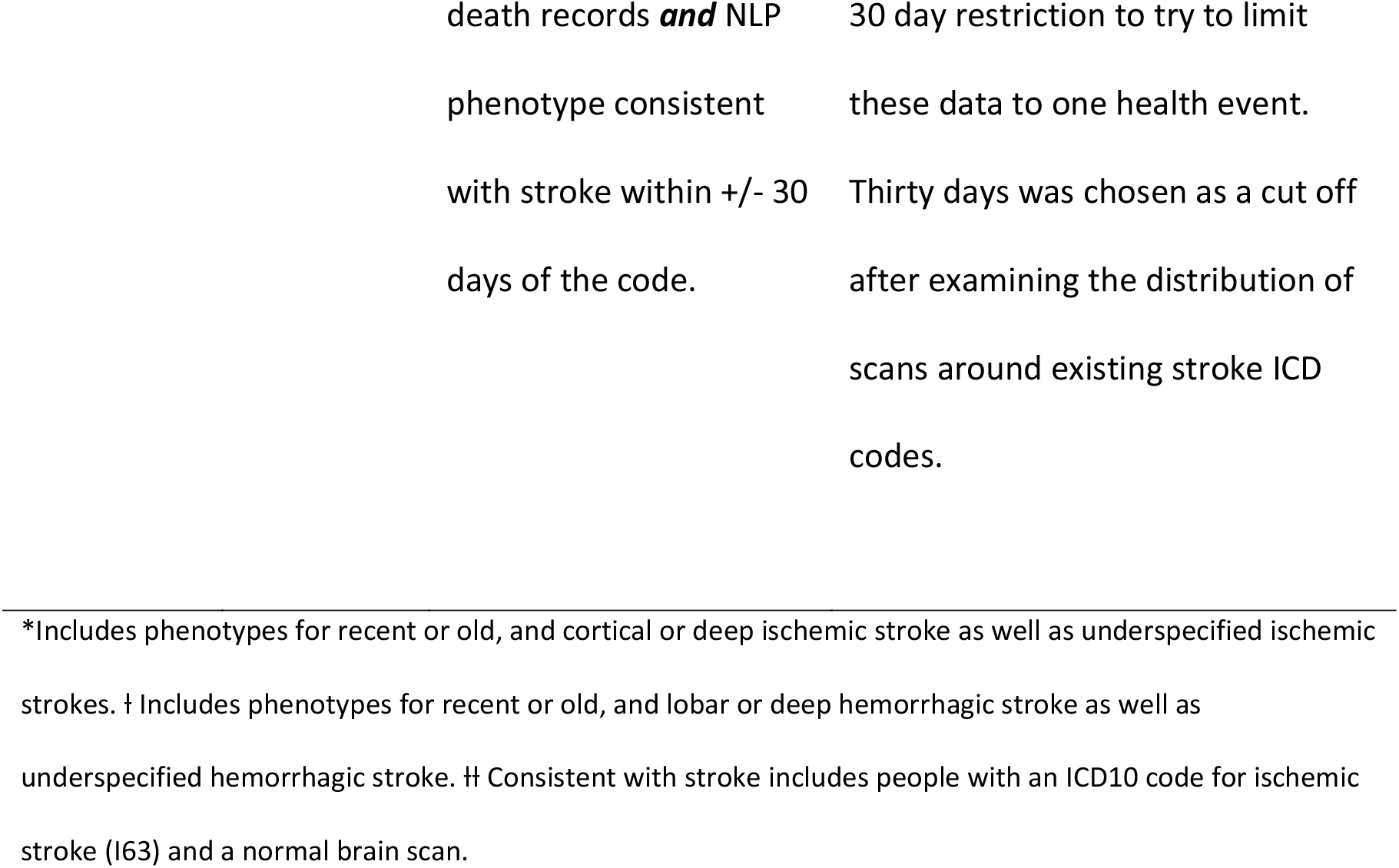
Definition of phenotype models

### Statistical analysis

First, we compared the baseline characteristics of participants with and without hypertension. Second, we looked at the brain imaging report metadata and brain phenotypes identified by EdIE-R. Lastly, we performed the following analyses for each of the five phenotype models:

- A calculation of crude and age-standardised incidence rate for all-stroke defined by each stroke phenotype model. We considered cases identified by ICD-10 codes to be acute cases. For the NLP data we included cases classified as “recent” and excluded any cases that were classified by EdIE-R as being “old” stroke only. Direct age standardisation was performed to the European Standard Population.^16^
- A Cox proportional hazards regression analysis to examine the relationship between baseline hypertension and the risk of all-stroke during follow-up (adjusted for age, sex smoking, BMI and history of existing diabetes, stroke or heart disease). In keeping with the above analysis, we only included NLP cases classified as being “recent” stroke, as we could not establish when an “old” stroke would have occurred Data was right censored for: people who withdrew from the GS study; died during follow up; at 2016 (31/12/16) for Tayside and Fife health boards and the other health boards at end of 2020 (31/12/20).
- Measurement of agreement between ICD-10 code and NLP stroke phenotype. We created a decision tree to assign a final phenotype for each stroke case when using data from both sources (Supplementary Table 1).

### Data cleaning and preparation

R Studio Version 1.4.1106 was used for all data cleaning, preparation, linkage and analyses.^17^

### Ethics

GS has ethical approval for the SFHS study (reference number 05/S1401/89) and 21CGH study (reference number 06/S1401/27) and both studies are now part of a Research Tissue Bank (reference 20-ES-0021).

## Results

### Dataset characteristics

After excluding participants with missing baseline data or who were not from our five selected health boards, there were 19,026 participants with a median age of 50 years, 11,375 (59.8%) were women, and 33% had hypertension at baseline. The characteristics of the participants with and without baseline hypertension are included in Supplementary Table 2. Our final study imaging dataset (Supplementary Figure 1) contained 3493 scans from 1938 participants (≥1CT: 1,056; ≥1 MRI: 513; both CT and MRI: 369 (Table 2)). From this dataset of brain imaging reports, NLP identified 1037 participants with ≥1 brain phenotype and 901 participants with no brain phenotype (Table 3). Stroke was identified in 534 participants: 319 with NLP alone, 59 with ICD-10 codes alone and 156 with both ICD-10 codes and an NLP report ‘consistent’ with stroke (this includes people with an ICD10 code for ischemic stroke (I63) and a normal brain scan; n=8). Of the cases with an NLP phenotype, there were 248 cases that had a phenotype for “recent” stroke.

**Table 2.** Characteristics of the Generation Scotland brain imaging report dataset (3493 scan reports)

|  |  | **Total**<br>**(n)** | **% of scanned**<br>**population** | **% of study**<br>**population** |
| --- | --- | --- | --- | --- |
| **Participants with at least one brain scan** |  | 1938 | 100 | 10.2 |
| **Participants with no brain scan** |  | 17088 | - | 89.8 |
|  |  |  |  |  |
| **Brain scan modality** | CT | 1056 | 54.5 | 5.6 |
|  | MRI | 513 | 26.5 | 2.7 |
|  | Both | 369 | 19.0 | 1.9 |
|  |  |  |  |  |
| **Number of scans per**<br>**person (n=3735 scans)** | 1 | 1254 | 64.7 | 6.6 |
|  | 2 | 391 | 20.2 | 2.1 |
|  | 3 | 116 | 6.0 | 0.6 |
|  | 4 | 59 | 3.0 | 0.3 |
|  | ≥5 | 118 | 6.1 | 0.6 |
|  |  |  |  |  |
| **Year of earliest scan** | 2006-08 | 70 | 3.6 | 0.4 |
|  | 2009-11 | 479 | 24.7 | 2.5 |
|  | 2012-14 | 466 | 24.0 | 2.4 |
|  | 2015-17 | 488 | 25.2 | 2.6 |
|  | 2018-20 | 435 | 22.4 | 2.3 |

CT: Computerised tomography; MRI magnetic resonance imaging

**Table 3.** Phenotypes identified by NLP of brain scan reports (per participant)

| **NLP output** | **Number of participants** | **% of scanned population** | **% of study population** |
| --- | --- | --- | --- |
| No phenotype on any scans | 901 | 46.5 | 4.73 |
| One or more phenotypes identified on scans | 1037 | 53.5 | 5.45 |
| **Phenotypes\*** |  |  |  |
| Small vessel disease | 568 | 29.3 | 2.99 |
| Atrophy | 467 | 24.1 | 2.45 |
| Any ischemic stroke | 370 | 19.1 | 1.94 |
| Any tumour | 176 | 9.08 | 0.93 |
| Hemorrhagic stroke | 101 | 5.21 | 0.53 |
| Stroke underspecified | 68 | 3.51 | 0.36 |
| Subdural haematoma | 54 | 2.79 | 0.28 |
| Subarachnoid hemorrhage | 53 | 2.73 | 0.28 |
| Microbleed | 19 | 0.98 | 0.10 |
| Hemorrhagic transformation | 19 | 0.98 | 0.10 |

\*People may have one or more phenotype identified and the categories are not mutually exclusive

### Testing stroke phenotype models

The results of the three analyses to test our phenotype models were as follows:

#### Incidence of all stroke

Table 4 shows the incidence of all-stroke calculated for the five stroke phenotype models. Age standardised rates were lowest for the phenotype based on ICD-10 stroke code and NLP stroke phenotype within +/- 30 days of the code (0.57 per 1000 person/years) and highest for the phenotype based on ICD-10 stroke code and/or NLP stroke (2.06 per 1000 person/years).

**Table 4.**
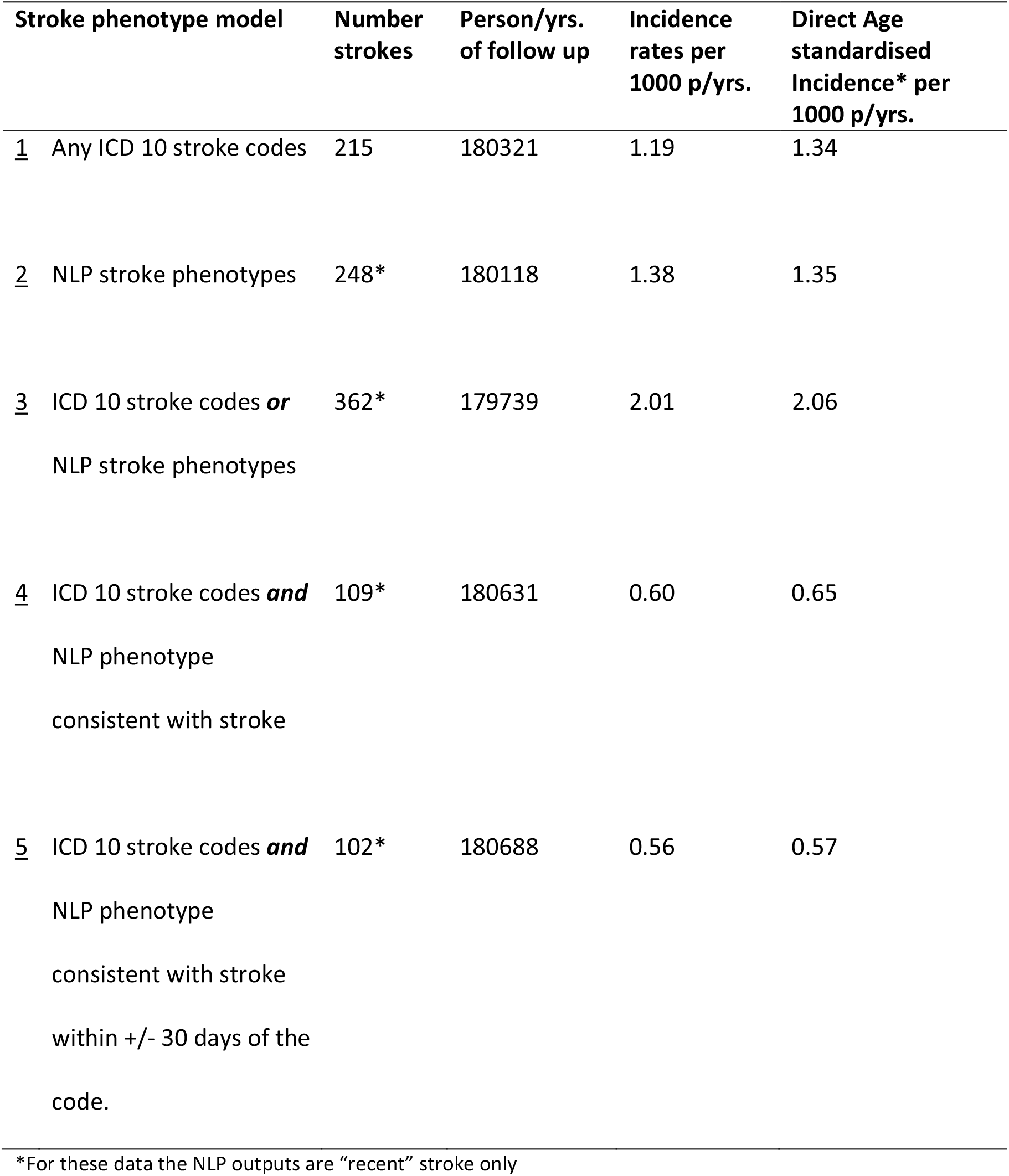
Incidence of stroke across the five stroke phenotypes

#### Risk factor validation

Figure 1 shows the estimated hazard ratios for hypertension for the five stroke phenotype models that were all relatively similar. The association between a diagnosis of hypertension at baseline and stroke was weakest when stroke was measured with the most inclusive phenotype (ICD-10 stroke code and/or NLP stroke phenotype, n=362, aHR 1.44 (95%CI [1.15-1.79] p = 0.0012)), and with the NLP stroke phenotype alone (n=248, aHR of 1.47 (95%CI [1.12-1.92] p= 0.0048). The remaining phenotypes (Phenotypes 1, 4, 5) identified fewer cases of stroke: ICD-10 stroke code alone n=215, ICD-10 stroke code and NLP stroke phenotype n=109 and ICD-10 stroke code and NLP stroke phenotype within +/- 30 days of the code n= 102. However, these phenotypes demonstrated a stronger association between hypertension and risk of all stroke: Phenotype 1 aHR: 1.57 (95%CI [1.18-2.10] p= 0.0021), Phenotype 4 aHR: 1.81 (95%CI [1.20-2.72] p=0.0046), and Phenotype 5 aHR: 1.75 (95%CI [1.15-2.67] p=0.0091). The aHR for these phenotypes including all NLP stroke cases (“recent” and “old”) are included for reference in supplementary Table 3.

**Figure 1.**
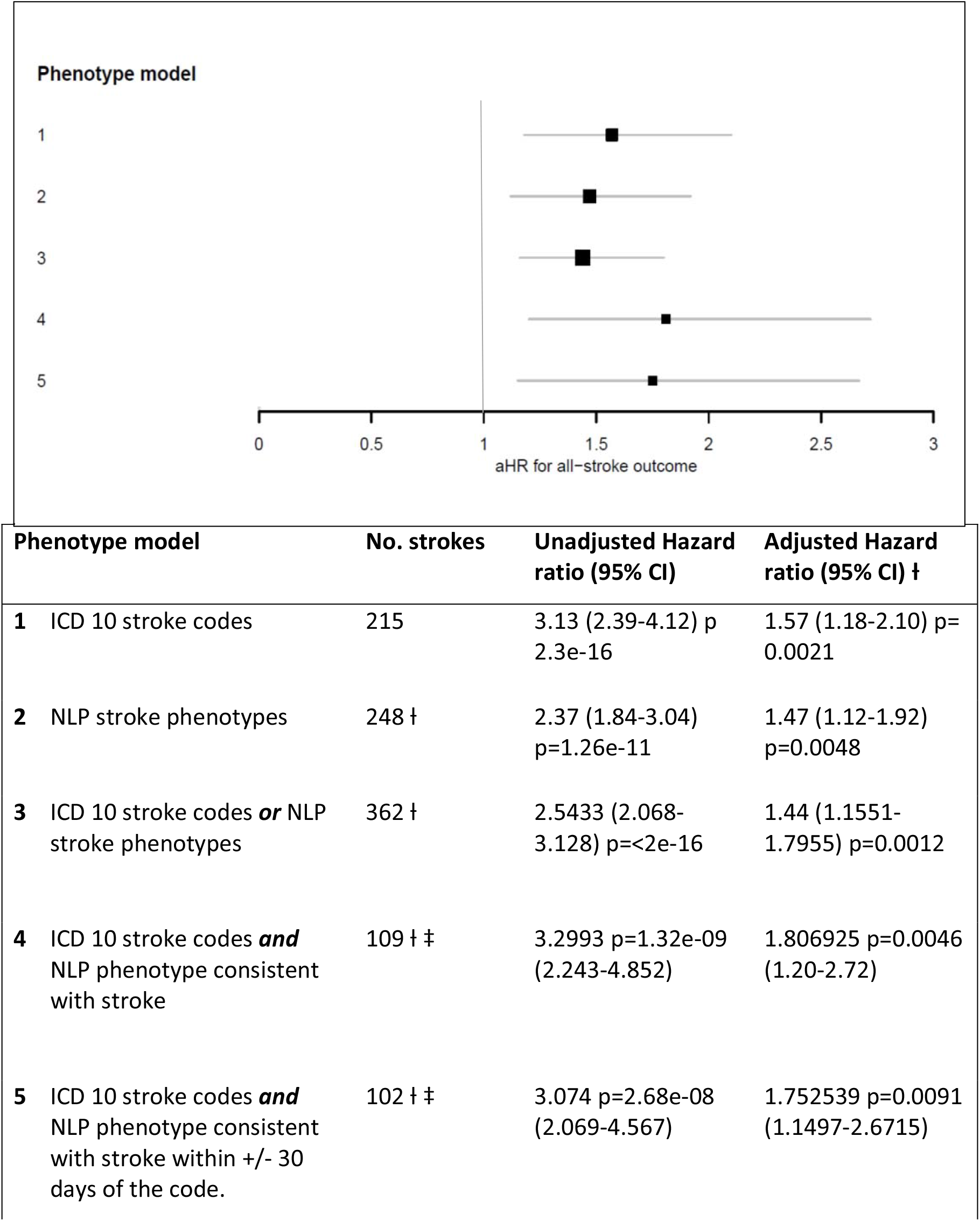
Hazard ratios for the association of baseline hypertension* and stroke outcome according to different stroke phenotypes *hypertension measured at baseline (defined as Systolic BP >140 mmHg and/or diastolic BP >90mmHg l For these data the NLP outputs are “recent” stroke only ‡ adjusted for bmi, smoking, sex, age, and history of existing diabetes, stroke or heart disease.

#### Analysis of stroke sub-type

Among people with ICD-10 stroke codes, 57/215 (26.5%) were coded as unspecified stroke (I64). The NLP phenotypes classified 42/467 (9.2%) cases as underspecified stroke. In 148 participants with both an ICD-10 code for stroke and an NLP phenotype for stroke, almost all cases could be assigned a stroke subtype (6/148, 4.0% unspecified; 118/148, 79.7% ischaemic stroke; and 24/148, 16.2% hemorrhagic stroke) (Figure 2). Consequently, adding NLP data lowered the number with no stroke subtyping in the coded dataset from 26.5% to 13.5% (29/215), and the overall dataset had 12% with no stroke subtype (65/534).

**Figure 2:**
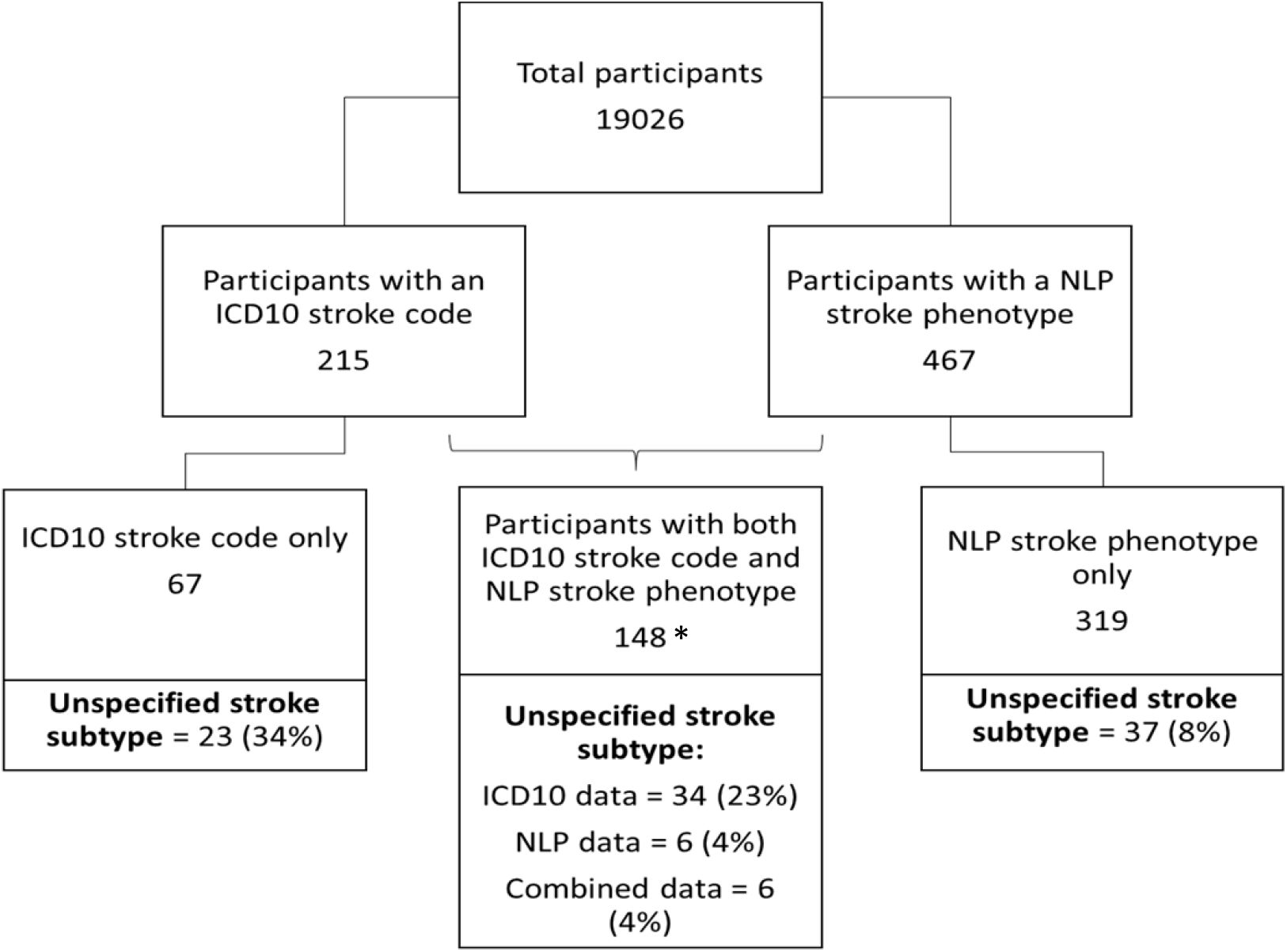
percentage of data with stroke sub-type unspecified according to data source * This doesn’t include the 8 cases which were consistent with ischemic stroke (i.e. an ischemic ICD10 code and normal brain scan report).

## Discussion

We used NLP to identify stroke phenotypes from routine clinical brain scan reports in a population-based cohort study. We tested five phenotype models with different combinations of NLP phenotypes and ICD-10 coded hospitalisation data, and tested these by estimating: the incidence of all-stroke; the size of association of hypertension with all-stroke; and the proportion of coded strokes that had no associated subtype. Where it is not possible to directly examine narrative EHR (as in this case), these methods provide indirect evidence of the phenotype validity.

We assumed greater misclassification of stroke would weaken the association between hypertension and stroke. The different stroke phenotype models had very modest differences in the size of the association between hypertension and all stroke. The aHR in this study lie within the range of positive associations demonstrated in existing studies, which vary according to factors such as the age of the population studied,^18, 19^ and geographical variation. ^20^ The HR of ischaemic stroke related to a systolic blood pressure of 144-159mmHg was found to be 1.64 (1.10-2.43) in a cohort study of middle aged men in South Wales and South West England.^21^ In an older cohort in England (aged over 65years and including both men and women) the HR of all-stroke related to a history of hypertension was 1.68 (1.35-2.09).^19^ A meta-analysis of risk factors for all-stroke in Western and Asian countries found a pooled HR for hypertension of 1.79 (1.39-2.30).^20^ The fact that all five phenotypes have the magnitude of association that we could expect to see between hypertension and stroke, provides added confidence in the validity of our ascertainment of stroke cases by ICD-10 codes, NLP output and combinations of these data.

The calculated standardised incidence of all-stroke was similar using ICD-10 coded data only or NLP phenotypes only, and higher when using the presence of any ICD-10 code or NLP phenotype. These results lie within previous estimates of stroke incidence within Europe at the start of the 21^st^ century ranging from 0.95 to 2.9 per 1000 per year (age-adjusted to the ESP).^22^ However, stroke incidence rates also vary geographically within Europe and are changing over time, and vary substantially by the method of ascertainment.^22^ Figures reported within the UK for the period of our study (approximately 2006 onwards) were from 0.9 to 1.14 per 1000 population.^23^ For Scotland specifically, estimated incidence rates calculated from hospital discharge data (age and sex adjusted to ESP) were 1.80 cases per 1000 population in 2012/13 and 1.76 per 1000 in 2021/22 (Figure 6), although these estimates include ICD-10 codes for Transient Ischaemic attack which were not included in our study.^24^

Using NLP data certainly improved detection of stroke cases, identifying 319 cases of stroke that had no corresponding ICD-10 codes. These cases may have been scanned as out-patients without ever being admitted to a hospital setting due to stroke, and are likely to represent less severe cases and may have been asymptomatic. NLP data also considerably improved the completeness of subtyping information. For cases with both coded and NLP data, the ICD-10 stroke codes had a much higher proportion where a subtype could not be assigned (23% compared with 4%). Having access to both data sources and using our decision tree (Supplementary Table 1) enabled us to determine a stroke subtype for a high proportion of the cases in the overall dataset (12% with no stroke subtype/88% subtyped). Subtype information is important for epidemiological research, including the exploration of risk factor associations and examining the effect of preventive and therapeutic interventions that will vary by subtype, due to their differing aetiologies. Additionally, NLP data offers more granularity in determining whether the reported strokes were “recent” or “old”, information used by our decision tree, and about their location within the brain, which may also be valuable information to identify cohorts for research studies.

In a subpopulation of the UK Biobank (UKB) cohort (N= 17,249)^1^ ICD-10 codes similarly identified a high proportion (42%) of stroke cases as unspecified stroke type. In a subsequent study, NLP combined with applied clinical knowledge was used on the imaging reports of the stroke cases identified by ICD-10 codes and they found that automated methods could assign a stroke type in 72% of cases. The authors concluded that for future research a combined approach using both coded data and NLP approaches is a feasible way to improve the accuracy of identifying stroke subtypes.^5^

Our study added to existing work as we applied NLP to a population-based cohort with brain image reports across five health boards and identified stroke cases with NLP (both old and recent strokes) that did not feature in the coded data. We also combined coded and NLP data into five different phenotype models and tested how these perform.

Much of the development of phenotype definitions, to date, uses differing data sources (e.g. ICD codes, Read codes, SNOMED codes, and free-text), with detailed decision processes. These descriptive algorithms can be difficult to transfer between EHR or health systems. ^10^ It can therefore be unclear how data sources are interacting and the influence this has on the composition of resultant definitions. The recent move towards creating computable phenotypes establishes an underlying authoring architecture ^25^ designed to provide a standard representation and to enable portability of phenotype definitions. In our study we attempted to unpack the contribution of NLP data to stroke phenotype definitions, with this in mind, to determine where NLP data has added value and to consider the best way to combine it with coded data. We hope that our findings, including the development of the decision tree, will assist in the addition of NLP data to computable phenotypes for stroke.

## Limitations

Limitations to this study were that: GS did not have complete baseline information on all participants; we were unable to obtain brain image reports from all health boards with GS participants; we did not have access to primary care records which may contain data on the results of out-patient brain scans; and we were not able to validate all of our stroke cases (code or NLP data) against a gold standard of physician adjudication of the complete EHR, but relied on risk factor validation. However, considerable clinician annotation was undertaken in the process of developing EdIE-R ^14^ and the clinical annotation that we carried out on the GS data as part of this study suggested that it was also performing well for this cohort.^15^

## Conclusion

We applied NLP to the radiology reports of all clinical brain scans carried out for GS participants to determine whether NLP data could improve the definition of stroke phenotypes over the use of coded data alone. The addition of NLP data to the phenotypes had the advantage of identifying more cases of stroke than coded data alone and was also advantageous in providing a more granular data source which enabled much more accurate subtyping of stroke. These findings provide valuable information on these two data sources and how they can be combined, and could assist in incorporating NLP data into a computable stroke phenotype. Further work is necessary to continue to develop this computable phenotype and to test it on other cohorts.

## Data Availability

The dataset Generation Scotland used in this study can be applied for via https://www.ed.ac.uk/generation-scotland/for-researchers/access.

## Non-standard Abbreviations and Acronyms

EHR: Electronic Health Record
GS: Generation Scotland
ICD 10: International classification of disease 10
NLP: Natural language processing
SMR: Scottish morbidity record
21CGH: GS 21st Century Genetic Health study
SFHS: GS Scottish Family Health Study

## Acknowledgments

This research was supported the Chief Scientist Office, The Alan Turing Institute, MRC, HDR-UK and Generation Scotland.

## Sources of Funding

- CSO
- BA, AC, and CG have been supported by The Alan Turing Institute via Turing Fellowships (BA, CG) and Turing project funding (ESPRC Grant EP/N510129/1).
- BA, CG, AMM and HCW were funded by MRC Mental Health Data Pathfinder Award (MRC-MCPC17209).
- ELB, MI and AMM are supported by the Wellcome Trust 220857/Z/20/Z.
- AMM and MI are supported by DATAMIND UKRI award MR/W014386/1
- ED was supported by the Alzheimer’s Society.
- Generation Scotland received core support from the Chief Scientist Office of the Scottish Government Health Directorates [CZD/16/6] and the Scottish Funding Council [HR03006] and is currently supported by the Wellcome Trust [216767/Z/19/Z].

## Disclosures

None

## Notes

### Competing Interest Statement

The authors have declared no competing interest.

